# Real-world uptake and outcomes of family screening in adults with thoracic aortopathy: a retrospective cohort study

**DOI:** 10.64898/2026.06.23.26356390

**Authors:** Maggie M. Pickard, Grace C. Potts, Morag C. Brown, Daniel J. Belliveau, Leslie Marcotte, Shanna Foster, John A. Sullivan, Christine R. Herman, Jeremy R. Wood, Kara Matheson, S. Gabrielle Horne

## Abstract

**Background:** Thoracic aortopathy is a disorder with genetic influence usually presenting in adulthood for which family screening is potentially desirable. Family screening is recommended, but the predictors of a positive screen and real-world pickup rates are unknown.

**Methods:** This was a retrospective cohort of 1022 probands (first affected family member identified) with thoracic aortopathy and one or more features suggestive of a genetic etiology, and their presenting family members, assessed in a cardiac clinic (2009–2024). Imaging and genetic testing were employed in family screening. The prespecified outcomes were uptake and pickup rate of family screening, and proband and family member specific characteristics that predicted a positive family screen.

**Results:** Among probands, 43.5% had one or more family member screened, with an average of 3 relatives per successful proband. 27.6% of family members screened positive. A pre-existing family history of aortopathy was the only variable predicting a higher incidence rate for positive family screen (p = 0.0003). Age of presentation < 60 was not predictive. For family members, extravascular features (p < 0.0001), closer relation to the proband (p < 0.02), male sex (p < 0.0001) and older age (p< 0.0001) all predicted a positive screen. Family members were eight times more likely to screen positive through imaging as compared to genetic testing. Probands with a genetic diagnosis of Marfan and Loeys Dietz syndromes accounted for only 4% of the total.

**Conclusions:** Proband-initiated family screening for thoracic aortopathy has a high yield of affected individuals, even among older probands.

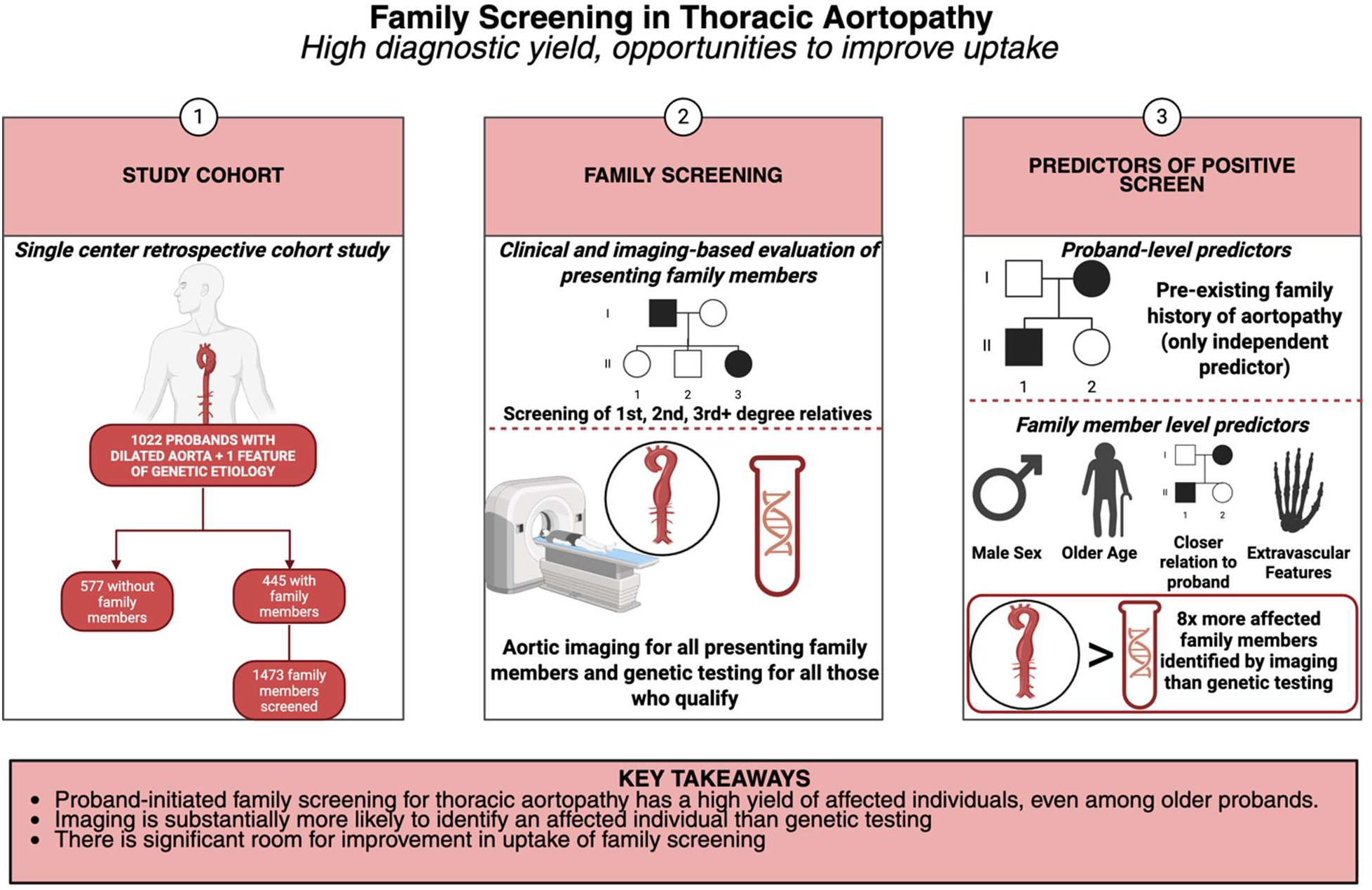

**Clinical perspective:**

- All patients with a dilated thoracic aorta, should be counselled on the benefits of family screening. Age over 60 at presentation and the absence of extravascular (syndromic) features in a proband (first affected family member) does not reduce the likelihood of a positive family screen.
- In families with known dissections or sudden death, family screening should be pursued beyond first degree relatives.
- An imaging first strategy for family screening is advised, rather than waiting for proband genetic testing results, given that eight times as many family members are picked up through aortic imaging as genetic testing.

## Introduction

There is strong evidence supporting genetic triggers for thoracic aortic aneurysms and dissection in subsets of affected patients^1–4^. Because thoracic aortic dissections have a high mortality^5–7^ and identification of thoracic aortopathy before dissection allows for elective repair to prevent this outcome, identification of early disease is advantageous. Some individuals with genetically triggered thoracic aortopathy have extravascular features and identifiable causative genetic variants, such as those associated with Marfan and Loeys Dietz syndromes^3^, others have causative genetic variants that are non-syndromic^3^, others have bicuspid aortic valve associated thoracic aortopathy, and still others have no currently identifiable causative variant but a familial pattern of thoracic aortopathy^8^. Because of the heterogeneity of clinical profiles among patients with genetically triggered thoracic aortopathy, it is unclear what proportion of patients with thoracic aortopathy have genetically triggered disease and what clinical factors predict a genetic trigger.

The outcomes of family screening have not been evaluated beyond small pilot studies employing classic cascade screening in subsets of patients with thoracic aortopathy^9,10^. There has been no systematic evaluation of proband or family member clinical features that predict a positive family screen in heterogeneous cohorts. Furthermore, cascade family screening do not track usual models of care in cardiac/cardiovascular clinics where most patients with thoracic aortopathy are typically assessed and managed. This has implications for the uptake of family screening as well as the applicability of data derived from cascade screening cohorts to broader patient populations.

Current best practice, as reflected in the guidelines, consists of family screening of first-degree relatives only, where an individual is suspected of having genetic thoracic aortopathy^11^. In these guidelines, it is assumed that age over 60 or the absence of extravascular features points to sporadic aortic disease^11^. Data is needed to guide the implementation of family screening, balancing responsible resource utilization with the opportunity to identify presymptomatic disease.

We sought to evaluate family screening for thoracic aortopathy in heterogeneous cohort of patients in a real-world cardiac/cardiovascular context, rather than cascade screening context. Specifically, we examined the uptake and outcomes of patient-initiated family screening in a large specialist cardiac clinic with a broad referral base, as well as the proband and family member clinical features predictive of a positive screen. The prespecified outcomes were (1) uptake and pickup rate of family screening with imaging and genetic testing, (2) proband and family member specific characteristics that predicted a positive family screen, defined as aortic dilatation, a bicuspid aortic valve, or a mid-sized artery aneurysm or dissection.

## Methods

### 2.1 Patient population

Health records of 8000 patients attending the Maritime Connective Tissue Clinic in Halifax, Canada between 2009 and 2024 were retrospectively reviewed. This clinic has a broad regional referral base from primary, secondary and tertiary care providers, and offers diagnostic assessments and care to patients and families at risk of genetic vasculopathy. We identified probands (the first affected family member known to the clinic) presenting with thoracic aortopathy and at least one feature of a genetic etiology (see below), as well as their family members referred to the clinic through proband-initiated family screening.

### 2.2 Inclusion and exclusion criteria

The inclusion of probands in this study required:

1. Patient is the first member of their family presenting to the clinic
2. Patient has either had a previous thoracic aortic dissection or has thoracic aortic dilatation, measured by CT angiography or MR angiography at either:
3. Aortic root measured at the level of mid-sinuses
4. Or ascending aorta, measured at the level of the pulmonary arteries
5. Patient has ≥ 1 feature suggestive of a genetic etiology
6. age of onset aortopathy aged <60 years (assuming dilatation rate of 1mm/year)
7. two or more extravascular features of genetic aortopathy, consisting of: craniofacial features (dolichocephaly, malar hypoplasia, narrow or high arched palate, broad or bifid uvula, retrognathia), scoliosis, kyphosis, pectus excavatum or carinatum, pes planus or foot deformity, protucio acetabulum or congenital hip dislocation, joint hypermobility, dural ectasia, abnormal striae, translucent or hyperextensible skin, atrophic scarring, unprovoked varicose veins, lens dislocation, retinal detachment, iris cysts, inguinal or abdominal hernias.
8. family history of aneurysm or vascular rupture/dissection
9. bicuspid aortic valve
10. midsized artery dissection or aneurysm There were no exclusion criteria.

### 2.3 Proband-initiated family screening

All probands with thoracic aortopathy and more than one risk factor for genetic etiology were educated on the potential autosomal dominant transmission of their aortopathy by the clinic nurse practitioner and/or physician as per clinic protocol. They were then provided with a letter that would allow relatives to access screening via the clinic. Interested family members were to bring this letter to a primary care practitioner to initiate referral. The letter included a unique identifier linking the family member(s) to the proband was included in the referral. The clinic thereby maintained kindred-based record keeping. All family members aged 16 and over were eligible for screening. Family members who screened negative at their initial assessment were offered re-imaging at 5-year intervals, unless genetic testing eliminated them from risk of transmission.

### 2.4 Data collection

Each patient assessed had a documented multigenerational pedigree, a personal medical history, cardiovascular examination and examination for extravascular features, echocardiogram and in those over age 30, a CT or MRI of the aorta. Data collected from probands’ and family members’ charts included demographics, history of hypertension, pre-existing family history of aortopathy, presence of extravascular features, distribution of aortic/arterial aneurysms and/or dissections from CT or MRI and aortic valve morphology from echocardiography. In addition, data was extracted as to whether they had been offered and undergone genetic testing, and the results. For family members, their relationship to the proband and the closest known affected family member was documented, as well as their status of follow up in the clinic.

### 2.5 Genetic testing

Clinic protocols prescribed that probands be offered referral to the regional genetics service for genetic testing if they met any of the following criteria: age of onset of aortopathy <50 years (assuming growth 1mm/year), marked extravascular features, or strong family history (typically aortic dissection or multiple suspicious sudden deaths without autopsy). Some patients were retested over time as the number of testable genes increased. When a pathogenic variant was identified in a proband, their family members were offered testing.

### 2.6 Positive family screen

A positive family screen from imaging was defined as one or more of: a definitively dilated aortic root or ascending aorta, as guided by a published nomogram^12^, a bicuspid aortic valve, or the presence of a mid-sized artery aneurysm or dissection identified on CT or MRI. A positive family screen from genetic testing was defined as a pathogenic variant identified in both the proband and a family member.

### 2.7 Ethical considerations

The use of de-identified patient data from clinical records for this research was approved by the Nova Scotia Health Research Ethics Board.

### 2.8 Statistical analysis

All analyses were performed using SAS STAT 15.1 version 9.4 (SAS Institute, Cary, N.C.). A two-sided P value of <0.05 was the threshold for statistical significance unless otherwise stated. Descriptive statistics were reported as counts and percentages for categorical variables, means ± standard deviation for normally distributed continuous variables, and medians and interquartile ranges for non–normally distributed continuous variables. A multivariable model was performed including all pre-selected variables. Incidence rate ratios are reported with 95% CI.

## Results

### 3.1 Proband clinical characteristics

Of the probands identified with thoracic aortopathy, 94.4% met one or more of our inclusion criteria. There were 1022 included (Figure 1). Their clinical characteristics are shown in Table 1. They ranged in age from 16 to 84. Of the probands with thoracic aortopathy, 26.9% had onset of thoracic aortopathy at age>60 years.

**Figure 1:**
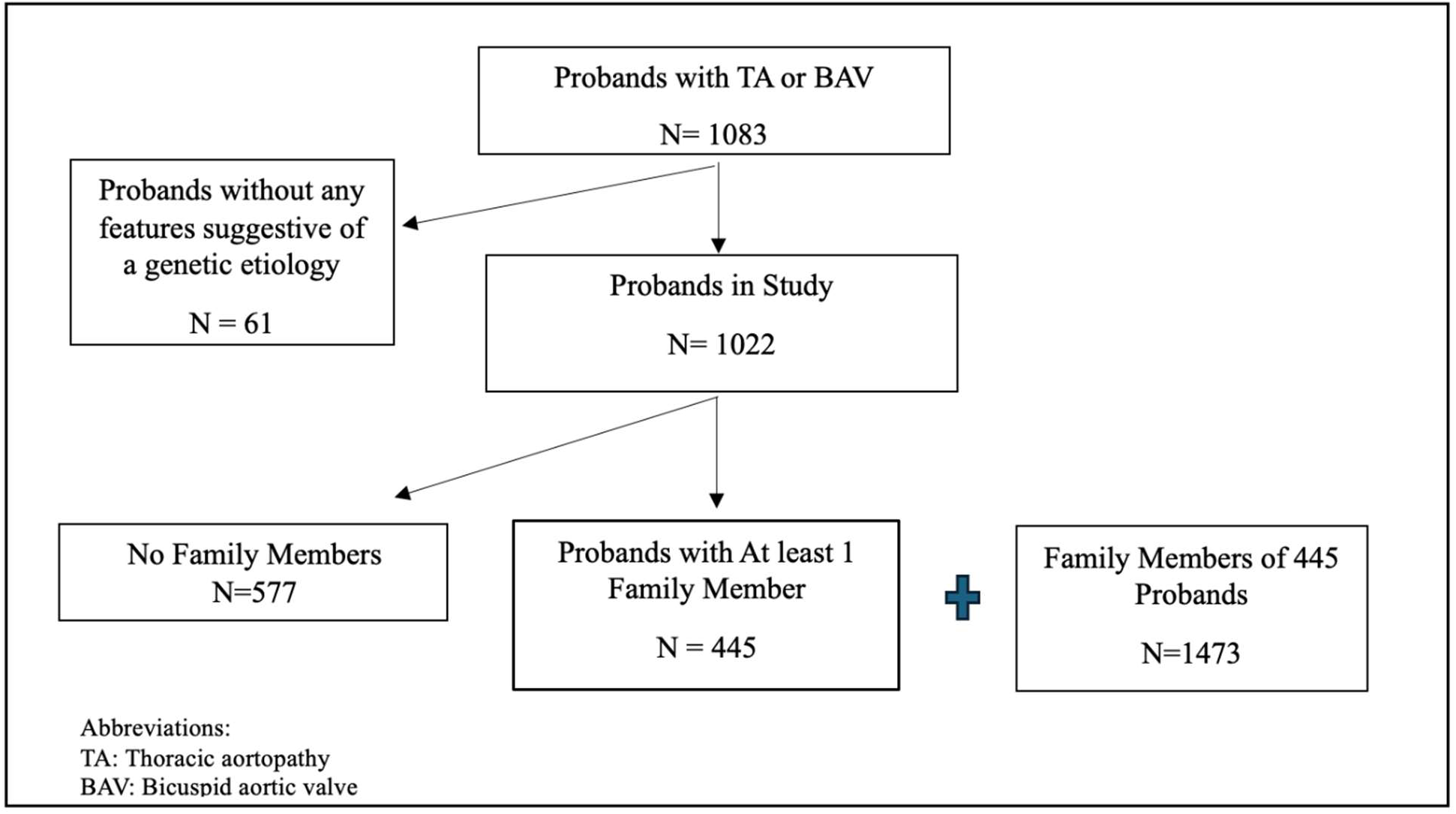
Study inclusion metrics. Flow diagram representing study inclusion and chart review results conducted at the Maritime Connective Tissue Clinic.

**Table 1:** Proband characteristics. Clinical characteristics of probands that visited the Maritime Connective Tissue Clinic between January 2008 and June 2024 that fit the inclusion criteria for the current study (N = 1022).

| **Proband characteristic** | **Total (N = 1022)** |
| --- | --- |
| *Mean age at presentation (SD)* | 51.1 (13.8) |
| *Median age at presentation (IQR)* | 53.2 (42.8, 60.6) |
| *Age at presentation < 60, n (%)* | 747 (73.1%) |
| *Female, n (%)* | 293 (28.7%) |
| *History of hypertension, n (%)* | 430 (42.1%) |
| *Presence of thoracic aortopathy, n (%)* | 974 (95.3%) |
| *Bicuspid valve, n (%)* | 239 (23.4%) |
| *Mid-sized artery aneurysm/dissection, n (%)* | 56 (5.5%) |
| *Prior aortic dissection, n (%)* | 86 (8.4%) |
| *Prior aortic surgery, n (%)* | 189 (18.5%) |
| *Presence of extravascular features, n (%)* | 606 (59.4%) |
| *Self-reported family history of aortopathy, n (%)* | 570 (55.8%) |
| *Self-reported family history of aortic dissection, n (%)* | 102 (10%) |

### 3.2 Family screening outcomes

Of the probands, 43.5% had one or more family members present to the clinic for family screening. Where family screening was successfully initiated, an average of 3.31 (SD 4.26) family members presented for each proband. The total number of family members ranged from 1-38. Of the 179 probands who had at least 3 family members present for screening, 70% had one or more family members screen positive. Having at least 3 family members screened was associated with at least 1 family member screening positive (p < 0.0001).

The clinical characteristics of the 1473 family members are described in Table 2. In total, 27.6% of family members screened positive. Of the 406 family members with a positive screen, 77.7% were first-degree relatives of the closest affected relative, 18.1% were second-degree, and 4.2% were third degree or more distant relatives. Of proband first degree relatives, 32% screened positive, compared to 22% of the more distant relatives.

**Table 2:**
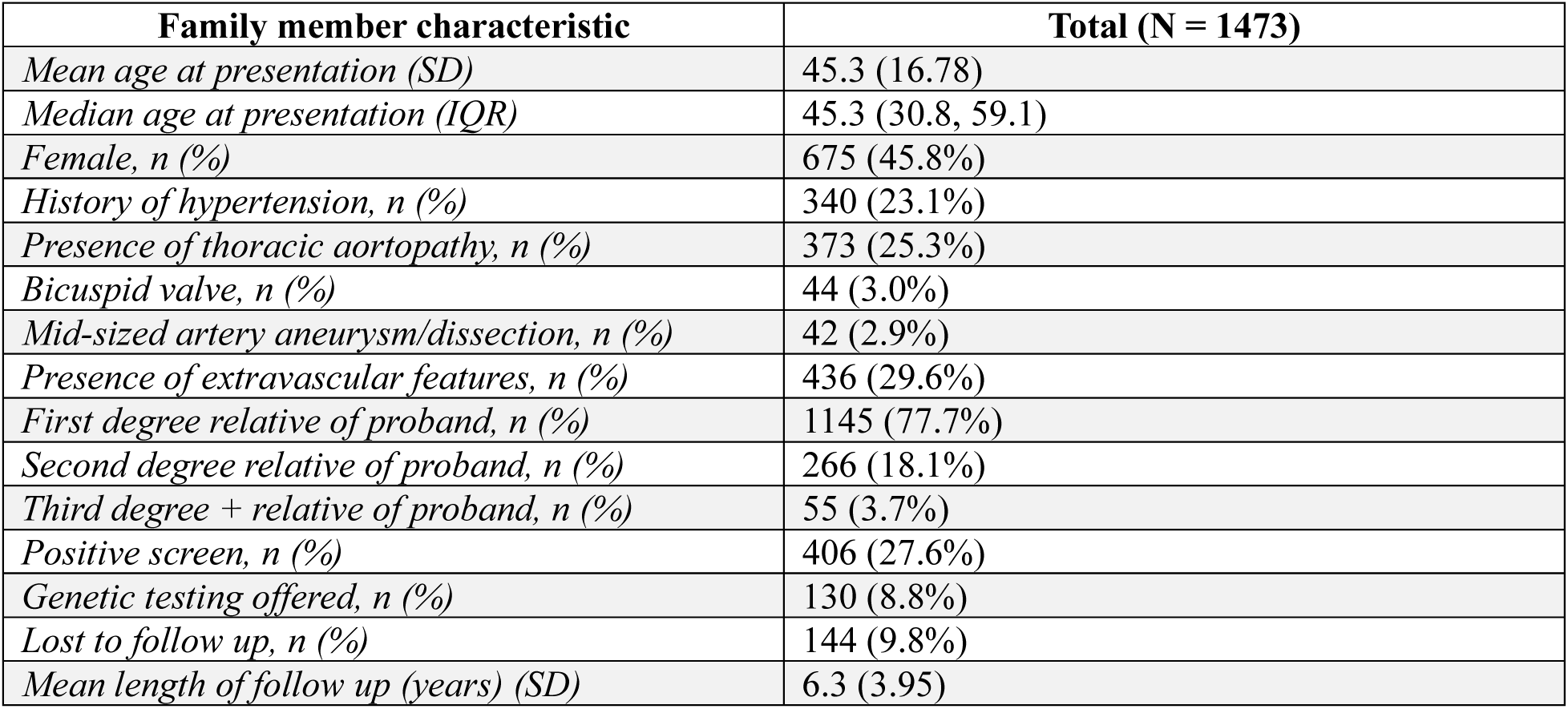
Family member characteristics. Characteristics of family members that presented to the Maritime Connective Tissue Clinic for aortic screening (N = 1473).

| **Family member characteristic** | **Total (N = 1473)** |
| --- | --- |
| *Mean age at presentation (SD)* | 45.3 (16.78) |
| *Median age at presentation (IQR)* | 45.3 (30.8, 59.1) |
| *Female, n (%)* | 675 (45.8%) |
| *History of hypertension, n (%)* | 340 (23.1%) |
| *Presence of thoracic aortopathy, n (%)* | 373 (25.3%) |
| *Bicuspid valve, n (%)* | 44 (3.0%) |
| *Mid-sized artery aneurysm/dissection, n (%)* | 42 (2.9%) |
| *Presence of extravascular features, n (%)* | 436 (29.6%) |
| *First degree relative of proband, n (%)* | 1145 (77.7%) |
| *Second degree relative of proband, n (%)* | 266 (18.1%) |
| *Third degree + relative of proband, n (%)* | 55 (3.7%) |
| *Positive screen, n (%)* | 406 (27.6%) |
| *Genetic testing offered, n (%)* | 130 (8.8%) |
| *Lost to follow up, n (%)* | 144 (9.8%) |
| *Mean length of follow up (years) (SD)* | 6.3 (3.95) |

### 3.3 Proband and family member specific predictors of a positive Screen

In univariate analysis of proband characteristics, adjusted for the number of family members presenting for each proband, pre-existing family history of aortopathy was the only variable that increased the odds of a positive family screen. The results of multivariate analysis of proband characteristics are summarized in Figure 2(A). After further adjusting for sex, age at presentation < 60 years and the presence of extravascular features, if a proband has a pre-existing family history of thoracic aortopathy, the incident rate of a screen positive family member was 1.60 times that for a proband with no pre-existing family history (p=.0003).

**Figure 2:**
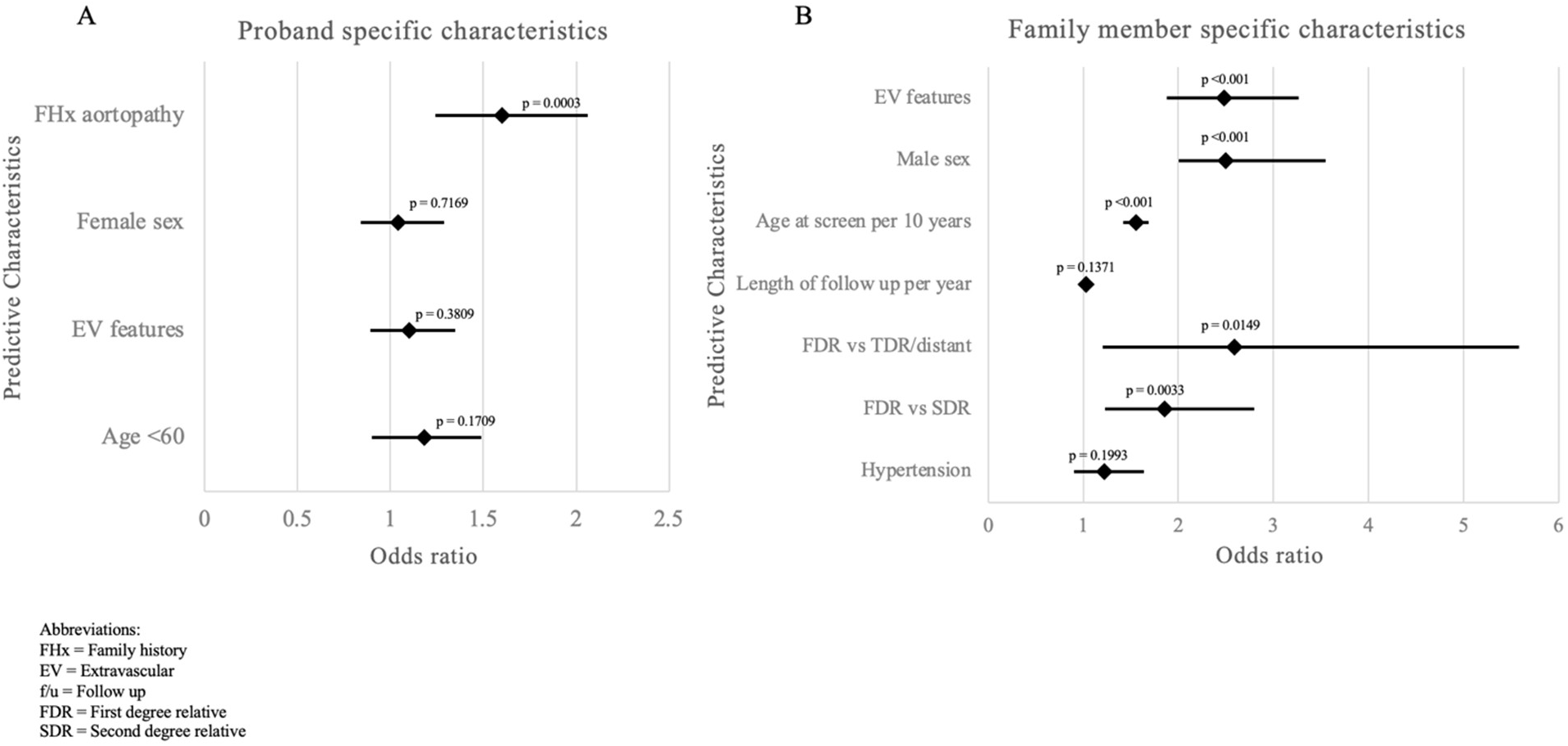
Predictors of a positive screen. **A)** Multivariate analysis of proband specific characteristics that predict whether a proband will have family members screen positive. **B)** Multivariate analysis of family member specific characteristics that predict whether a family member will screen positive.

Characteristics of the family members that significantly predicted whether they would have a positive screen in univariate analysis were history of hypertension, closer relationship to proband, age at first screen, male sex, and extravascular features. When accounting for other variables in multivariate analyses, significant family member characteristics predicting a positive screen shifted slightly, as summarized in Figure 2(B). The odds of a positive screen for a family member who is a first degree relative of a proband is 2.59 times that of a third degree or distant relative (p = 0.0149), and 1.85 times that of a second degree relative (p = 0.0033). For each 10-year increase in age at time of screening, odds of a positive screen increase 55.06% (p<0.0001). Male family members have 2.47 times greater odds of screening positive than a female (p < 0.001). Finally, if a family member has extravascular features, they are 2.47 times more likely to screen positive compared to a family member without (p < 0.0001).

### 3.4 Genetic testing

372 probands were offered genetic testing. Of these, 54% had an age of onset <50 years and 53.2% had a pre-existing family history of aortopathy or aortic dissection. The proportion offered testing solely based on marked extravascular features was 15.6%.

Of note, 27.4% of probands (14.3% of females and 32.4% of males) offered genetic testing either declined to complete or were lost to genetic follow up. Of the 270 who completed testing, 70 had pathogenetic variants. 61.4% of these pathogenic variants were in genes associated with Marfan’s syndrome (MFS) fibrillin-1 gene (FBN1) and 23.2% were in genes associated with Loey-Dietz syndrome (LDS). The remaining 14.4% of pathogenic variants were in smooth muscle and collagen genes (Figure 3). Gene positive probands make up only 4.8% of all probands included in the study, 18.8% of those offered genetic testing and 25.9% of those probands that underwent genetic testing. Probands with a genetic diagnosis of Marfan syndrome and Loeys Dietz syndrome combined accounted for only 4% of the total probands in this study.

**Figure 3:**
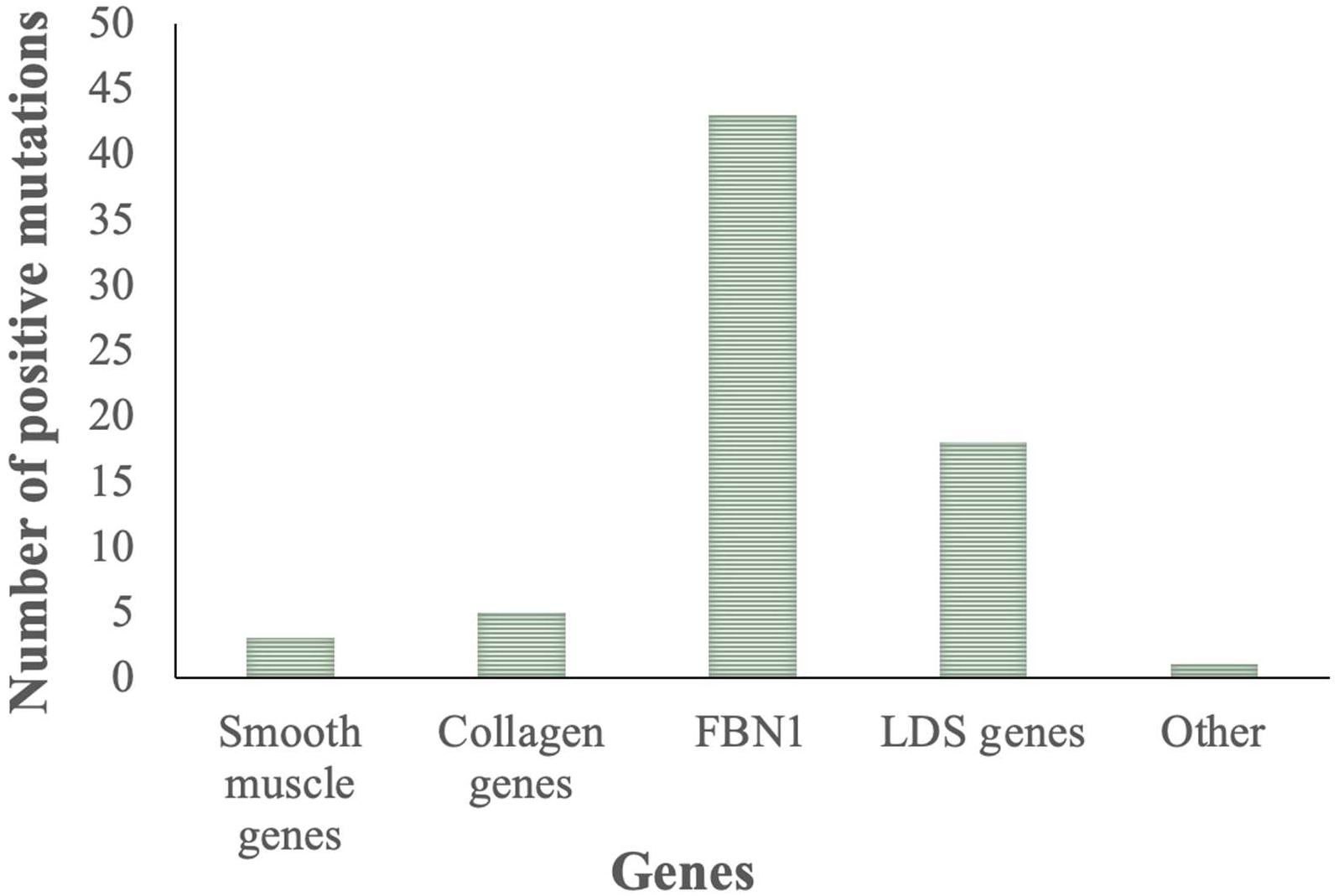
Distribution of genes in positive mutations. Distribution of the genes in this cohort of the 70 probands who were found to be genetically positive

105 family members related to these 70 gene positive probands completed genetic testing, and 51 had pathogenic variants. This represents 3.5% of the total family members screened in this study, as compared to the 28% who screened positive with imaging.

Univariate and multivariate analyses were performed to determine which characteristics predicted a pathogenetic variant being identified in probands who completed genetic testing. The presence of extravascular features, pre-existing family history of aortopathy, and female sex predicted a positive genetic test. Age at presentation <60 was not predictive. When correcting for other variables in multivariate analyses, these results was unchanged (Figure 4).

**Figure 4:**
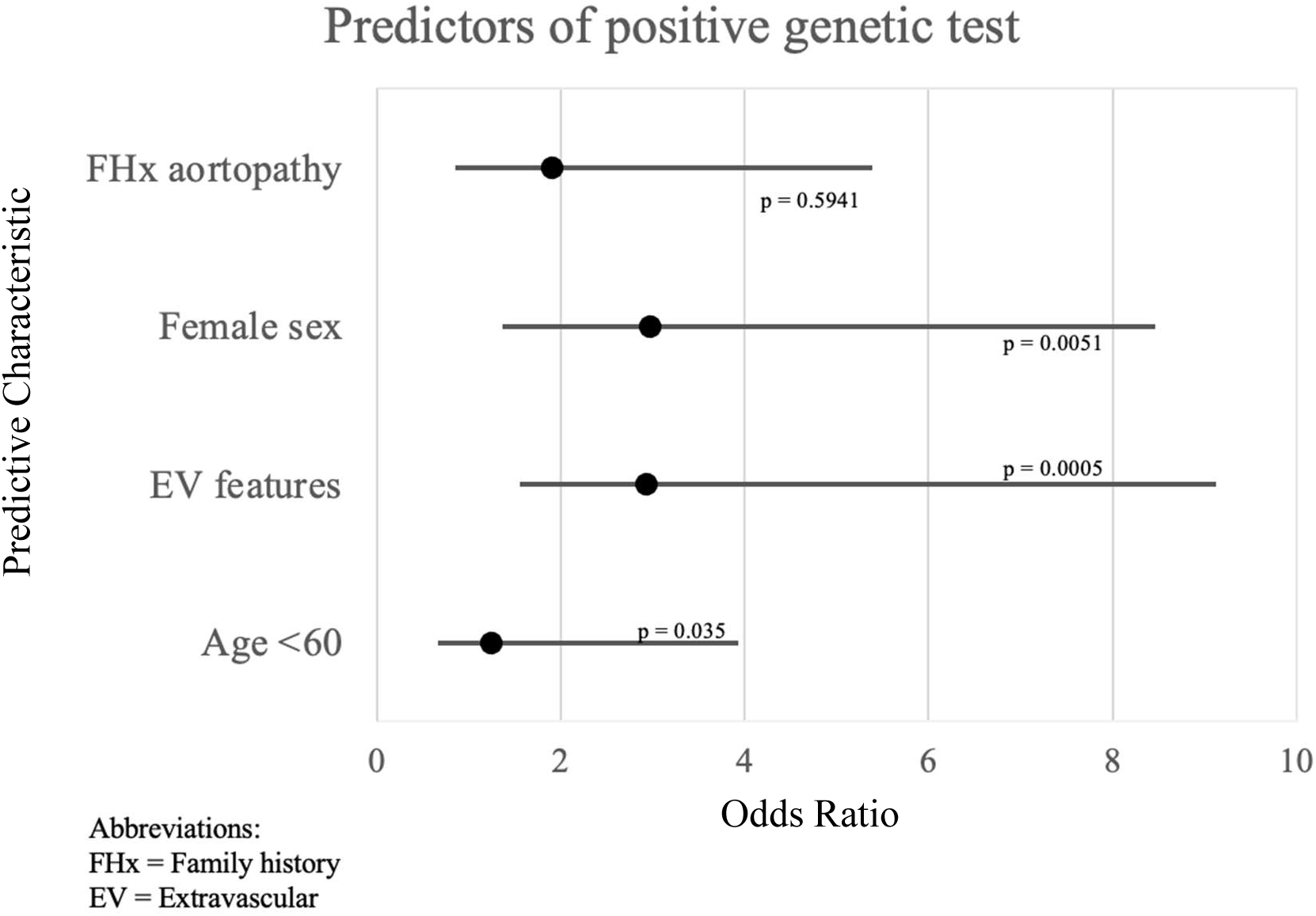
Predictors of appositive genetic test. Multivariate analysis of proband specific characteristics that predict whether a proband will have a positive genetic test. Female sex, age less than 60, and extravascular features were all statistically significant predictors of probands having a genetic mutation found via genetic testing (p < 0.05).

## Discussion

We have shown that for a heterogeneous cohort of patients with thoracic aortopathy, proband-initiated family screening yielded 43% of probands having one or more family members present for screening, 27% of these screened positive, with 25% having dilated aortic root or ascending aortas. Importantly, only a small minority of patients included in this study were gene positive and less than 5% of probands had a diagnosis of Marfan or Loeys Dietz syndrome.

Hence this is the first highly generalizable cohort described for the purposes of family screening. Among probands with thoracic aortopathy, those with age of onset < 60 years and extravascular features did not have a higher chance of a positive family screen. However, among family members, extravascular features did predict a positive screen. As expected, older family members and those most closely related to the proband were more likely to have a positive screen. Despite restricting genetic testing to those with the strongest clinical evidence of a genetic etiology, only a quarter of those probands who completed genetic testing had pathogenic variants identified. Additionally, over a quarter of probands referred for genetic testing did not follow through. Due to a combination of these two factors, family members were eight times as likely to screen positive through imaging as through genetic testing.

Genetic thoracic aortopathies are typically transmitted in an autosomal dominant pattern, with some data indicating either reduced penetrance or variable expression^13,14^. However there remains some question as to what proportion of thoracic aortopathy is genetic^19^. In addition to evidence of monogenetic mendelian inheritance, there is also evidence of thoracic aortopathy being inherited as a complex trait with gene loci associated with increased risk in older adults^15^. Our results are interpretable as supporting a predominantly autosomal dominant pattern of inheritance with variable penetrance, and greater aortic phenotypic expression at an older age. Our data does not support using onset or presentation before age 60, as a cutoff to separate a genetic etiology from sporadic thoracic aortopathy in probands, as has previously been proposed^11^. Nor does our data support using the absence of extravascular features in a proband as a rationale to withhold family screening. Further study is warranted to determine the outcomes of family screening in thoracic aortopathy presenting in the elderly.

### 4.1 Outcomes of patient-initiated screening

Ours is the largest proband cohort reported to date; 10 times larger than a 2022 study by Abbasciano et al evaluating the feasibility of cascade screening in non-syndromic probands^9^.

While a 2016 study of cascade screening reported similar total numbers to our study, with 2385 patients, there were only 270 probands and were again limited to non-syndromic thoracic aortic disease^10^.

Classic cascade screening begins with finding an affected proband then extending testing to at-risk biologic relatives, starting with first degree relatives, and typically directed through a genetic counsellor^5^. Indirect cascade screening differs as contact with family members is arranged through the proband, instead of a genetic counsellor. Our study used proband-initiated family screening, where probands were educated and provided with a letter to pass on to any family members to initiate screening at the clinic, and any presenting family members were accepted for screening. This screening strategy likely tracks typical real-world models of care for most patients who are assessed and managed in cardiac/cardiovascular clinics. Proband-initiated family screening removes any requirement for a health care professional to orchestrate the process. This human resource savings should be balanced against the potential for probands being ineffective at initiating family screening, and the possibility of imaging resources being expended on distant relatives at minimal aortic risk.

Our uptake from patient-initiated family screening is on par with cascade screening in the REST study conducted by Abbasciano et al., where uptake of family screening was 34%, despite professional oversight^9^. In the 2016 Australian study, 58% of identified relatives offered cascade screening came forward^10^, and they screened 2.15 family members per probands with a total of 270 probands who gave consent for family screening. By comparison, our study screened 1.44 family members per proband counting the total 1022 probands in our study. However, if the 445 probands in our study that did bring forward family members is considered analogous to the 270 consenting probands in the Australian study, our uptake from proband-initiated screening is larger at 3.3 family members per proband. We therefore conclude that the uptake we report from proband-initiated family screening is comparable to that of cascade screening in a similar patient population. The potential concern of distant relatives at minimal risk consuming resources was not seen in our study. Only 4% of presenting family members were third-degree or more distantly related to the closest affected person in their family.

Current guidelines only recommend screening first-degree relatives of affected individuals^11^. In our cohort, 32% of these screened positive, while of the remaining non-first-degree relatives, 22% had positive screens. We suspect probands used their education on extravascular features, acquired in their own clinical assessment, to identify more distant relatives at higher risk of being affected, thereby enriching the pickup rate for distant relatives. The pickup rate we report in non-first-degree relatives is sufficient that consideration should be given to screening more distant relatives, particularly where there is a family history of aortic dissection, given the potential consequences of a missed diagnosis.

Despite our yield being comparable to cascade screening, the majority of probands failed to initiate family screening, with over 80% of probands having less than 3 family members screened. Further study is needed to identify barriers to facilitating family screening. A 2020 Delphi exercise conducted on genetic thoracic aortopathy noted the importance of streamlined processes, coordinated interprofessional care and patient education^16^.

### 4.2 Proband and family member characteristics

Although genetic aortopathy is often described in binary terms as syndromic or non-syndromic, our patients presented on a spectrum from none to marked extravascular features. Importantly, the presence of extravascular features in probands did not predict a more penetrant pattern of inheritance. In contrast, when family members had extravascular features, they were 2.5 times more likely to screen positive than those without. The apparent paradox, where extravascular features were discriminating for family members but not probands, is likely because only a quarter of family members were affected but most probands had evidence of a genetic etiology. Because extravascular features are not always present in genetic aortopathy, their presence would not be discriminating in identifying affected individuals if most of the proband pool were affected.

For each 10-year increase in age of family members at time of screening, the odds of a positive screen are 55.1% greater. Longer length of follow up also significantly predicted increased odds of a positive screen. This aligns with the progressive nature of aortopathy and supports periodic repeat aortic imaging for family members of gene negative probands, with an initial negative imaging screen.

### 4.3 Genetic testing

Among our probands with positive genetic testing, the distribution of pathogenic variants among the genes known to trigger thoracic aortopathy was in line with the current literature^17^.

The criteria used for offering genetic testing were similar to those recommended in the current guidelines^11^, allowing for a time-lapse between age of onset and age of presentation with thoracic aortopathy. The large difference between the utility of imaging and genetic testing for family screening in our study suggests that an imaging first rather than genetic testing first approach should be adopted for family screening, particularly in clinical environments with significant wait times for testing.

Genetic testing required the proband to attend a further assessment appointment, and 27.4% did not follow through with this. Mainstreaming of genetic testing, as is done in some cancer screening programs^18,19,20^ may reduce this dropout. Further study is warranted to determine the outcomes of genetic testing of less stringently selected probands with thoracic aortopathy.

### 4.4 Sex differences

Probands were more likely to be male, and male family members were more likely to screen positive. Previous studies have reported a higher prevalence of aortopathy in men than women^21,22^ and our results point to increased penetrance in men. Unexpectedly, female sex was a predictor of a positive genetic test. We speculate that because female sex attenuates penetrance in aortopathy, those women with disease penetrant enough to meet criteria for genetic testing likely have variants in genes associated with more aggressive disease, more likely to have already been identified in this evolving field. This is supported by a 2022 study that shows average age of onset for thoracic aortopathy genes is younger than the average in the population^23^.

### 4.5 Limitations

The uptake of family screening was likely underestimated as children were not screened and undoubtedly some family members were screened outside the study, particularly those living at a distance. Probands who were tested in early calendar years, found to be gene negative and never re-tested, may have had pathogenic variants that could have been identified with contemporary aortic panels. Hence, we may have underestimated the pickup rate of genetic testing.

### 4.6 Conclusion

Proband-initiated family screening for thoracic aortopathy is effective, and high yield when imaging is used alongside genetic testing. Arbitrary age cut-offs and the absence of extravascular features do not exclude a genetic etiology in probands. The pickup rate in non-first-degree relatives is significant. Family members were eight times as likely to screen positive with imaging as with genetic testing in this real-world heterogeneous cohort. Delivery of scalable, pragmatic family screening models holds promise for improving outcomes of thoracic aortopathy.

## Data Availability

The data that supports the findings of this study are available from the corresponding author upon a reasonable request

## Non-standard Abbreviations and Acronyms

